# Is coagulation-protein consumption upon admission linked to COVID-19 severity and mortality?

**DOI:** 10.1101/2021.04.19.21255747

**Authors:** Francisco C. Ceballos, Pablo Ryan, Rafael Blancas Gómez-Casero, María Martin-Vicente, Erick Joan Vidal-Alcántara, Felipe Peréz-García, Sofía Bartolome, Juan Churruca-Sarasqueta, Ana Virseda-Berdices, Oscar Martínez-González, Oscar Brochado-Kith, Marta Rava, Carolina Vilches-Medkouri, Natalia Blanca-López, Ignacio Ramirez Martinez-Acitores, Patricia Moreira-Escriche, Carmen De Juan, Salvador Resino, Amanda Fernández-Rodríguez, María Ángeles Jiménez-Sousa

## Abstract

The link between coagulation system disorders and COVID-19 has not yet been fully elucidated. With the aim of evaluating the association of several coagulation proteins with COVID-19 severity and mortality, we performed a cross-sectional study in 134 patients classified according to the highest disease severity reached during the disease. We found higher levels of antithrombin, prothrombin, factor XI, factor XII and factor XIII in asymptomatic/mild and moderate COVID-19 patients than healthy individuals. Interestingly, decreased levels of antithrombin, factor XI, XII and XIII were observed in those patients who eventually developed severe illness. Additionally, survival models showed us that patients with lower levels of these coagulation proteins had an increased risk of death. In conclusion, COVID-19 provokes early increments of some specific coagulation proteins in most patients. However, lower levels of these proteins at diagnosis might “paradoxically” imply a higher risk of progression to severe disease and COVID-19-related mortality.

## INTRODUCTION

Coronavirus disease 2019 (COVID-19) is associated with a significant activation of the coagulation cascade. While thrombosis has been classically described in acute and chronic infections including respiratory diseases (1), thrombotic risk appears to be higher in COVID-19 (2). Consequently, thromboembolic complications are common in hospitalized patients, especially among those in intensive care units (ICU) (3). In this setting, several mechanisms of coagulation activation have been postulated (4) and large dynamic fluctuations in coagulation and fibrinolysis laboratory parameters have been described during disease course (5). Development of overt disseminated intravascular coagulation (DIC) seems to be rare and to follow a different pattern from other infection-derived DIC (5-7), but it has been reported in up to 71% of fatal cases as a late and ominous sign (8).

Additionally, both venous and arterial thrombotic events have been independently associated with mortality (9). Several haemostatic-system abnormalities such as thrombocytopenia, elevated D-dimer levels, prolonged prothrombin time (PT) or activated partial thromboplastin time (APTT), decreased factor V activity, hypofibrinogenemia and reduced levels of natural anticoagulants (e.g antithrombin) appear with increasing disease severity and are linked to death (6, 8) (10). However, the bidirectional relationship between SARS-CoV-2 and the coagulation system is still not completely understood (4). A predominant increase of D-dimer is typical, and its presence on admission has been repeatedly described as significantly higher in non-survivors (11) but scarce or no abnormalities in PT and APTT are usually found at disease onset (5, 12). To date, coagulation markers measured in the early phase of COVID-19 have evidenced a complex scenario and elucidation of the pathophysiology of immunothrombosis is evolving. Therefore, to continue unravelling the insights of COVID- 19-induced coagulopathies, we evaluate several coagulation proteins at an early stage of disease onset and their association with the highest disease severity and mortality.

## METHODS

### Design and study population

Cross-sectional study in 128 COVID-19 patients enrolled from March to September 2020 at three hospitals in Madrid (Infanta Leonor University Hospital, Aranjuez University Hospital and Príncipe de Asturias University Hospital). The study protocol was approved by the Ethics Committee of the Institute of Health Carlos III (PI 33_2020-v3) and the Ethics Committee of each hospital.

Patients were classified according to their highest severity grade during the course of COVID-19, (**Supplementary Figure 1**): 1) Severe: i) death during hospitalization, ii) ICU admission, iii) invasive mechanical ventilation; or iv) presence of bilateral pulmonary infiltrates, mechanical ventilation and oxygen saturation (Sat0_2_) ≤93%; 2) Moderate: the remaining hospitalized patients who did not fulfill severe COVID-19 criteria; 3) Asymptomatic/Mild (AM): individuals who had minor or no COVID-19 symptoms; 4) A control group of 16 pre-pandemic healthy controls without any known infection was included. The STROBE-ID checklist was used to strength the design and conduct the study.

### Clinical data and samples

Epidemiological, clinical, disease evolution data, as well as laboratory parameters such as PT, international normalized ratio (INR) and APTT were collected from clinical records using an electronic case report form (eCRF) which was built using REDCap (13). Plasma samples were obtained after centrifugation blood in EDTA tubes at hospital admission (median = 2 days, IQR = 4 days). Samples were processed at the National Center for Microbiology (Majadahonda), Institute of Health Carlos III (Madrid, Spain).

### Coagulation proteins

Coagulation proteins (antithrombin, prothrombin, factor XI, factor XII and factor XIII) were measured by ProcartaPlex Panel (Invitrogen) multiplex immunoassay by using a Luminex 200™ analyzer (Luminex Corporation, United States) according to the manufacturer’s specifications.

### Statistical Analysis

As outcome variables, the highest disease severity during COVID-19 infection and the mortality were considered. For descriptive data, differences between groups were tested using Chi^2^ or Fisher’s exact test with Monte Carlo simulated p-value for categorical and Kruskal-Wallis test for continuous variables. The association between coagulation proteins, measured in the first days of disease, and the severity was explored using generalized linear mixed models (GLMM) (See Supplementary material and methods for extended information). Survival time was defined as time between hospitalization and death, and individuals alive at 90 days were considered censored data. Survival curves were modelled using Kaplan Meier method and log rank test was performed to assess univariate differences in survival time according to coagulant proteins tertiles levels. Death risk was estimated with the Cox Proportional-Hazard and Aalen’s additive models. Age and sex were included as covariables. Two-sided tests were used for all statistical methods. Analyses were performed using the R 4.0.3 software.

## RESULTS

Patient’s characteristics are shown in **Table 1**. Coagulation proteins were mostly measured before therapy administration, and no associations were found with comorbidities, chronic medications or COVID-19 presenting (**Supplementary Table 1, Supplementary Figure 2**), showing no prior bias. Additionally, APTT and PT (INR) levels did not show differences among the three groups of COVID-19 (**Supplementary Figure 3)**.

**Table 1.** Patient’s characteristics. Patients were classified according to the highest disease severity reached during the COVID-19 evolution. Denominator indicates number of patients with available data. **Statistics**: Individual characteristics were summarized using standard descriptive statistics: mean ± standard deviation for continue variables and count (percentage) for categorical variables. Differences between groups were tested using Fisher’s exact test. No statistically significant association was found between covariates and disease severity with the exception of the following treatments: Cloroquine/hydroxycloroquine (Fisher’s exact test = 1.6e^-05^), Tocilizumab (Fisher’s exact test = 8.1e^-05^), corticosteroids (Fisher’s exact test = 5.7e^-08^), and the use of supplemental oxygen (Fisher’s exact test = 3.2.e^-07^). **Abbreviations**: NA, non-available ; BMI, body mass index; NSAIDs, Nonsteroidal anti- inflammatory drugs; ACE, Angiotensin-converting-enzyme; ARBs, angiotensin II inhibitors; HIV, human immunodeficiency virus; ICU, intensive care unit.

|  | COVID-19 Severity |  |  |  |
| --- | --- | --- | --- | --- |
|  | Healthy | Asymptomatic/Mild | Moderate | Severe |
| **Demographics** |  |  |  |  |
| N | 16 | 13 | 68 | 47 |
| Age | 58.8 ± 12.3 | 64.3 ± 18.1 | 61.1 ± 15.3 | 62.1 ± 18.1 |
| Gender (male) | 9/16 (56.2%) | 5/13 (38.4%) | 38/68 (55.8%) | 33/47 (70.2%) |
| BMI ≥ 25 | 8/11 (72.7%) | 2/12 (16.6%) | 11/68 (16.1%) | 13/47 (27.6%) |
| Smoke status (Yes) | NA | 2/13 (15.3%) | 3/68 (4.4%) | 3/47 (6.3%) |
| Former smoker | NA | 4/13 (30.7%) | 8/68 (11.7%) | 11/47 (23.4%) |
| **Comorbidities** |  |  |  |  |
| Hypertension | NA | 5/13 (38.4%) | 30/68 (44.1%) | 23/47 (48.9%) |
| Cardiopathy | NA | 4/13 (30.7%) | 12/68 (17.6%) | 8/47 (17.0%) |
| Chronic pulmonary disease | NA | 0/13 (0%) | 8/68 (11.7%) | 10/47 (21.2%) |
| Chronic kidney disease | NA | 2/13 (15.3%) | 4/68 (5.8%) | 9/47 (19.1%) |
| Chronic liver disease | NA | NA | 4/65 (6.2%) | 2/47 (4.2%) |
| Chronic neurological disease | NA | 0/13 (0%) | 9/67 (13.4%) | 8/47 (17.0%) |
| Neoplasia | NA | NA | 3/63 (4.8%) | 4/47 (8.5%) |
| Diabetes | NA | 2/13 (15.3%) | 10/68 (14.7%) | 11/47 (23.4%) |
| Chronic inflammatory disease | NA | NA | 2/63 (3.1%) | 5/47 (10.6%) |
| Autoimmune disease | NA | NA | 1/63 (1.5%) | 4/47 (8.5%) |
| **Therapy** |  |  |  |  |
| **Chronic medications** |  |  |  |  |
| NSAIDs | NA | 0/13 (0%) | 0/68 (0%) | 7/47 (14.9%) |
| ACE inhibitors | NA | 0/13 (0%) | 15/68 (22.3%) | 11/47 (23.4%) |
| ARBs | NA | 1/13 (7.6%) | 7/68 (10.4%) | 8/47 (17.0%) |
| Corticosteroids | NA | NA | 6/68 (9.5%) | 6/47 (12.8%) |
| HIV antiretrovirals | NA | NA | 1/68 (1.5%) | 1/47 (2.1%) |
| **Treatment** |  |  |  |  |
| Chloroquine and hidroxychloroquine | 0/16 (0%) | 0/13 (0%) | 62/63 (98.4%) | 33/47 (70.2%) |
| Tocilizumab | 0/16 (0%) | 0/13 (0%) | 10/67 (15.0%) | 20/47 (42.5%) |
| Corticosteroids | 0/16 (0%) | 0/13 (0%) | 26/67 (38.8%) | 40/47 (85.1%) |
| **COVID-19 related symptoms** |  |  |  |  |
| Dyspnoea | 0/16 (0%) | 0/13 (0%) | 43/68 (63.2%) | 41/47 (87.2%) |
| Cough | 0/16 (0%) | 0/13 (0%) | 48/63 (76.1%) | 34/47 (72.3%) |
| Headache | 0/16 (0%) | 0/13 (0%) | 24/63 (38.1%) | 10/47 (21.2%) |
| Diarrhea or abdominal pain | 0/16 (0%) | 0/13 (0%) | 33/63 (52.4%) | 19/47 (40.4%) |
| **Hospitalization** |  |  |  |  |
| Length of stay (days) | 0/16 (0%) | 0/13 (0%) | 10.7 ± 8.6 | 22.1 ± 15.6 |
| Maximum Temperature | NA | NA | 37.8 ± 0.8 | 38.2 ± 0.8 |
| Oxygen therapy | 0/16 (0%) | 0/13 (0%) | 39/68 (57.3%) | 47/47 (100%) |
| Non-Invasive mechanical ventilation | 0/16 (0%) | 0/13 (0%) | 3/67 (4.5%) | 20/47 (38.99%) |
| Invasive mechanical ventilation | 0/16 (0%) | 0/13 (0%) | 0/67 (0%) | 18/47 (38.2%) |
| Pulmonary infiltrates | 0/16 (0%) | 0/13 (0%) | 60/68 (88.2%) | 47/47 (100%) |
| **ICU** |  |  |  |  |
| ICU length of stay | 0 | 0 | 0 | 6.61 ± 13.5 |
| Death | 0/16 (0%) | 0/13 (0%) | 0/68 (0%) | 18/47 (38.3%) |

Distribution of antithrombin, prothrombin, factor XI, factor XII and factor XIII protein levels for each disease severity group is shown in **Figure 1A**. Statistical differences between severity groups are shown through the GLMM in **Figure 2**. AM, moderate and severe individuals had higher concentrations of coagulation proteins compared to healthy individuals. Also, we detected a significant reduction of factor XI, factor XII and factor XIII concentration in severe patients respect to moderate individuals (**Figure 2; Supplementary Table 2**). These pairwise differences between COVID-19 severity groups were also detected at individual protein level (**Figure 2; Supplementary Table 3**). Kaplan- Meier analyses showed that the patients with lower levels of the analysed proteins had an increased risk of death during hospitalization (**Figure 1B)**. For antithrombin, prothrombin, factor XI and XIII, this increase is more accentuated in men **(Supplementary Figure 4**). The effect of proteins levels on the survival is confirmed by the Cox proportional-hazard models and Aalen’s additive regression (**Supplementary Table 4**). Both models found a significant effect of the different coagulation proteins on the survival. These models found a negative correlation between the proteins’
s concentration and the time of survival (**Supplementary Table 4**). A reduction in the coagulation proteins concentration involves a higher risk of death, however, the effect size was slight for each individual protein (**Supplementary Table 4**). ROC curves showed that the addition of the coagulation proteins to the basic model composed by sex and age slightly improved the survival prediction although was not significant (**Supplementary Figure 5**).

**Figure 1.**
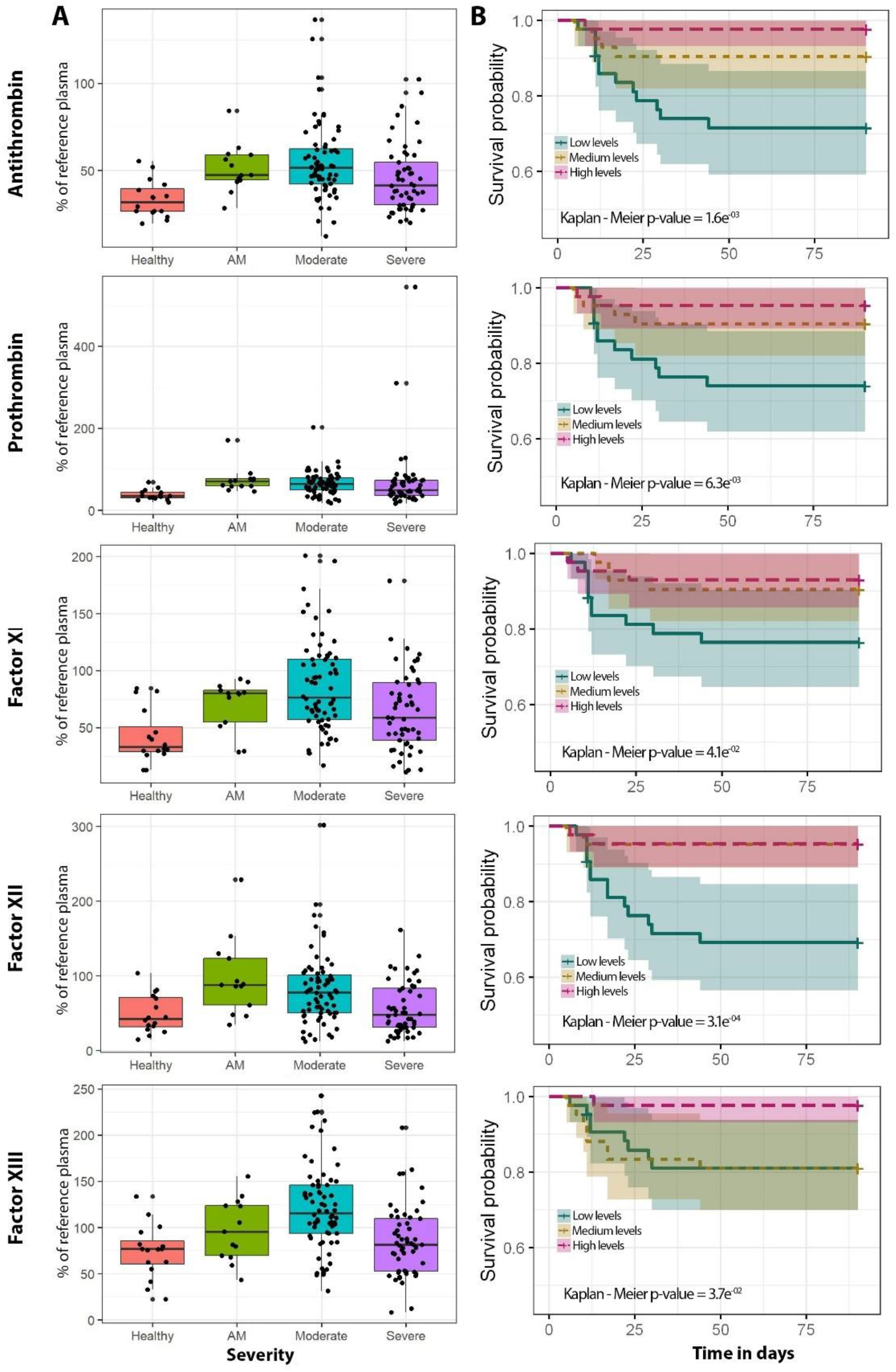
Coagulation proteins levels regarding COVID-19 severity and survival analysis for each of the coagulation proteins analysed in this study. **A**. Boxplots of healthy (purple, n=16), AM (blue, n=13), moderate (green, n=68) and severe (yellow, n=47) individuals. P-values from ordinal logistic regression (OLR) are shown in each boxplot. **B**. Kaplan-Meier plot. The cut-offs of coagulation proteins for the Kaplan-Meier plot were obtained using three quantiles to get low (blue), medium (yellow) and high (pink) percentage of the reference plasma. The specific categorical levels by using tertiles were as follows: Antithrombin: low [12.14 – 41.3], medium [41.3 – 54.1], high [54.1 – 136.7]. Prothrombin: low [16.3 – 48.7], medium [48.7 – 72.2], high [72.2 – 545]. Factor XI: low [10.8 – 56.4], medium [56.4 – 90.1], high [90.1 – 200.9]. Factor XII: low [11.7 – 47.8], medium [47.8 – 87.1], high [87.1 – 301.9]. Factor XIII: low [8.3 – 82.6), medium [82.6 – 118.3], high [118.3 – 242.7]. **Abbreviations:** AM, Asymptomatic/Mild patients; OLR, ordinal logistic regression.

**Figure 2.**
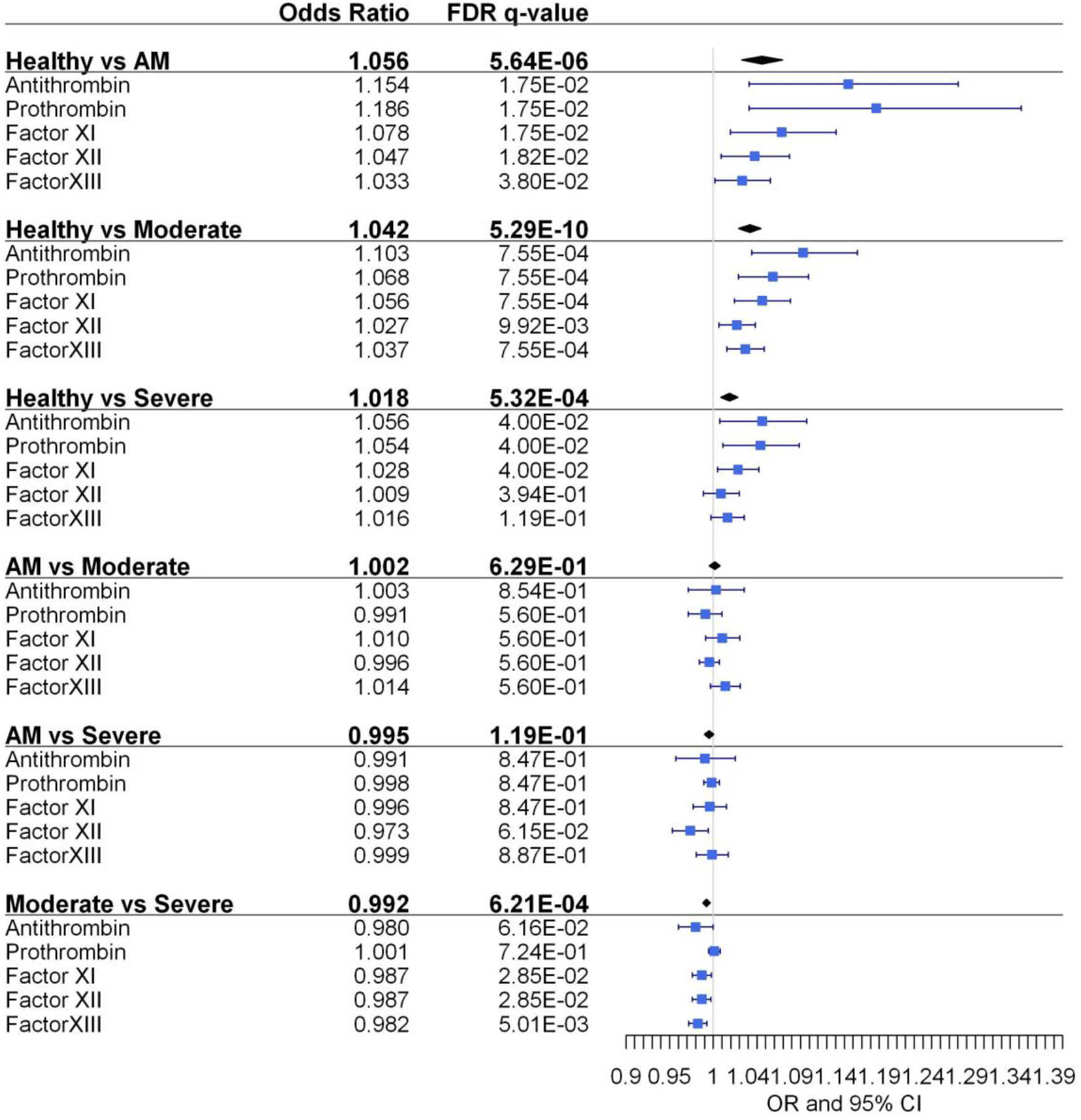
Pairwise association between coagulation protein levels and the different severity classes. Statistics: Pairwise comparisons between COVID-19 severity classes were obtained using a logistic mixed model where the protein was considered a random effect. (see Supplementary Material and Methods). Pairwise comparisons within each protein were obtained using multiple logistic regression analysis. The False Discovery Rate or FDR was used to cope with multiple testing, q-values are provided. **Abbreviations**: AM, Asymptomatic/Mild patients.

## DISCUSSION

Our study shows that coagulation protein levels are affected at the first stages of the COVID-19 infection and that those early changes already reflect disease severity in its acute phase. We report increased levels of antithrombin, prothrombin, factor XI, factor XII and factor XIII in AM and moderate patients, compared to healthy individuals. The increase in natural anticoagulant and procoagulant proteins in COVID-19 has been attributed to thromboinflammatory response caused by SARS-CoV-2, which provokes endotheliitis and increases the hepatic production of factors (14). These could explain the increases of clotting proteins found in our study even in early and mild stages. In contrast, significantly decreased levels of antithrombin, factor XI, XII and XIII were found at presentation in severe with respect to moderate patients. Therefore, an early reduced production or more likely increased consumption due to pulmonary or systemic coagulopathy of clotting proteins in severe cases could predict a poor prognosis. Results of the survival models are in accordance with previous studies (15) that show a consumption of coagulation proteins among COVID-19 non-survivors. Besides, it is important to note that this negative association was more pronounced in men.

Several limitations should be considered. First one is the limited sample size, of healthy and asymptomatic cases, which could have limited the possibility to find statistical significance differences in some comparisons. Second one is that we have analysed five coagulation protein; as coagulation cascade is extremely complex, further studies should consider additional factors to fully describe the COVID-19 effects over the entire coagulation cascade. However, these additional factors have been extensively studied, and we analysed those that were not previously addressed.

In conclusion, our results indicate that: 1) COVID-19 causes an early increase of some specific coagulation proteins such as antithrombin, prothrombin, contact factors and factor XIII in most patients, even in those who won’t suffer from clinically significant disease, suggesting that commonly elevated D-dimer levels are driven by an initial enhanced procoagulant state and not just by hyperfibrinolysis; 2) Although not reflected in routine tests such as PT and APTT, and despite common initial hyperfibrinogenemia, patients who will eventually advance to severe disease show early decreased levels of these anticoagulant and procoagulant markers, suggesting factor consumption, an these levels were associated with higher COVID-19 related mortality. Evolving investigations will allow us to better clarify the crosstalk between the immune and clotting systems in this pandemic disease.

## Data Availability

Data Availability on request.

## ACKNOWLEDGEMENTS

This study would not have been possible without the collaboration of all the patients, their families, medical and nursery staff, and data managers who have taken part in the project.

## ACCESS TO DATA

All data are available upon request to corresponding authors.

## CONTRIBUTION

Funding body: AFR and MAJS.

Study concept and design: AFR, MAJS and FCC

Patients’ selection and clinical data acquisition: PR, RBGC, MMV, AVB, OBK, FCC, FPG, OMG, CVM, NB, IRMA.

Sample preparation, and biomarker analysis: MMV, AVB, EJVA, SB, OBK.

Statistical analysis and interpretation of data: FCC, AFR, and MAJS.

Writing of the manuscript: FCC, AFR, and MAJS.

Critical revision of the manuscript for relevant intellectual content: PR, RBGC, FPG, JCZ, OMG, MR, PME, CJ, SR.

Supervision and visualization: AFR and MAJS.

All authors read and approved the final manuscript.

## SUPPLEMENTARY MATERIAL AND METHODS

### Statistical Analysis

We use a generalized linear mixed model (GLMM) to test the pairwise difference among disease severity classes by grouping the four coagulation proteins analysed in this study. We fit a model where the protein was considered a random effect:

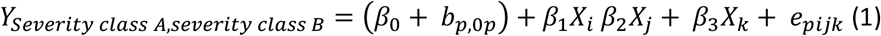

Where b_p,0p_ is the random effect of each coagulation protein, β_1_X_i_ is the fixed effect of the protein concentrations, β_2_X_j_ is the fixed effect of the age and β_3_X_k_ is the fixed effect of the sex. By using this model, we tested whether we were able to detect general effects between disease severity classes. We were also interested in study the behaviour each coagulation protein. Pairwise comparisons between disease severity classes, for each protein, were carried out using multivariable logistic regressions. Sex and age were included as covariables in the multivariable analysis.

Survival Cox Proportional-Hazard model’s goodness of fit was evaluated by the Harrel’s concordance index (C-index). This index ranges from 0 to 1 and the intuition behind it is that, if the risk model is good, patients who had shorter times-to-death should have higher risk scores. Values of C-index near 0.5 indicates that the risk score predictions are not better than chance in determining which individual with die first. Coagulation proteins discrimination capabilities were measured by the area under the receiver operating characteristic (ROC) curve.

**Supplementary Table 1.** Differences in the levels of coagulation proteins regarding discrete covariables.

|  | **Antithrombin** | **Prothrombin** | **Factor_XI** | **Factor_XII** | **Factor_XIII** |
| --- | --- | --- | --- | --- | --- |
| Sex | 0.996 | 0.933 | 0.933 | 0.971 | 0.623 |
| BMI>=25 | 0.832 | 0.683 | 0.683 | 0.799 | 0.623 |
| Smoker | 0.600 | 0.323 | 0.364 | 0.517 | 0.314 |
| Hypertension | 0.832 | 0.683 | 0.683 | 0.872 | 0.623 |
| Cardiopathy | 0.832 | 0.933 | 0.933 | 0.799 | 0.658 |
| Chronic pulmonary disease | 0.832 | 0.323 | 0.364 | 0.517 | 0.917 |
| Chronic kidney disease | 0.832 | 0.323 | 0.364 | 0.799 | 0.314 |
| Chronic liver disease | 0.947 | 0.683 | 0.683 | 0.872 | 0.658 |
| Chronic neurological disease | 0.832 | 0.933 | 0.933 | 0.872 | 0.623 |
| Neoplasia | 0.832 | 0.683 | 0.683 | 0.872 | 0.624 |
| Diabetes | 0.956 | 0.933 | 0.933 | 0.948 | 0.314 |
| Chronic inflammatory disease | 0.625 | 0.933 | 0.933 | 0.872 | 0.413 |
| Autoimmune disease | 0.996 | 0.933 | 0.933 | 0.872 | 0.643 |
| NSAIDs | 0.947 | 0.718 | 0.718 | 0.958 | 0.658 |
| ACE inhibitors | 0.947 | 0.683 | 0.683 | 0.932 | 0.658 |
| ARBs | 0.947 | 0.683 | 0.683 | 0.932 | 0.623 |
| Dyspnoea | 0.996 | 0.683 | 0.683 | 0.932 | 0.658 |
| Cough | 0.996 | 0.683 | 0.683 | 0.981 | 0.658 |
| Headache | 0.915 | 0.946 | 0.946 | 0.902 | 0.314 |
| Diarrhea or abdominal pain | 0.600 | 0.323 | 0.364 | 0.517 | 0.717 |

**Statistics:** Differences were evaluated by Kruskal Wallis rank sum test (False Discovery Rate (FDR) q-values for the Kruskal Wallis are shown). **Abbreviations:** BMI, body mass index; NSAIDs, Nonsteroidal anti-inflammatory drugs; ACE, Angiotensin-converting-enzyme; ARBs, angiotensin II inhibitors.

**Supplementary Table 2.** Pairwise differences between COVID-19 severity classes.

|  | OR | 2.5% | 97.5% | FDR q-value |
| --- | --- | --- | --- | --- |
| **Healthy vs AM** |  |  |  |  |
| Prot. Concentration | 1.056 | 1.033 | 1.080 | 5.64E-06 |
| Age | 0.946 | 0.918 | 0.976 | 4.12E-04 |
| Sex | 0.289 | 0.119 | 0.700 | 5.98E-03 |
| **Healthy vs Moderate** |  |  |  |  |
| Prot. Concentration | 1.042 | 1.029 | 1.055 | 5.29E-10 |
| Age | 0.983 | 0.965 | 1.001 | 7.07E-02 |
| Sex | 1.222 | 0.705 | 2.119 | 4.76E-01 |
| **Healthy vs Severe** |  |  |  |  |
| Prot. Concentration | 1.018 | 1.008 | 1.028 | 5.32E-04 |
| Age | 1.004 | 0.987 | 1.021 | 6.74E-01 |
| Sex | 2.428 | 1.420 | 4.151 | 1.19E-03 |
| **AM vs Moderate** |  |  |  |  |
| Prot. Concentration | 1.002 | 0.995 | 1.008 | 6.29E-01 |
| Age | 1.021 | 1.005 | 1.037 | 1.04E-02 |
| Sex | 3.181 | 1.785 | 5.670 | 8.65E-05 |
| **AM vs Severe** |  |  |  |  |
| Prot. Concentration | 0.995 | 0.990 | 1.001 | 1.19E-01 |
| Age | 1.018 | 1.002 | 1.034 | 2.72E-02 |
| Sex | 5.297 | 2.889 | 9.715 | 7.11E-08 |
| **Moderate vs Severe** |  |  |  |  |
| Prot. Concentration | 0.992 | 0.988 | 0.997 | 6.21E-04 |
| Age | 1.010 | 0.999 | 1.021 | 6.87E-02 |
| Sex | 1.918 | 1.335 | 2.755 | 4.26E-04 |

**Statistics:** A general linear mixed model (GLMM) was fit considering the coagulation protein a random effect. Protein concentration, age and sex were considered fixed effects. Odds ratio, 95% confidence intervals (lower boundary: 2.5%, upper boundary: 97.5%), and the False Discovery Rate (FDR) q-values are shown for every model.

**Supplementary Table 3.** Association between coagulation proteins and COVID-19 severity.

|  | OR | 2.5% | 97.5% | FDR q-value |
| --- | --- | --- | --- | --- |
| **Healthy vs AM** |  |  |  |  |
| Antithrombin | 1.155 | 1.041 | 1.280 | 1.75E-02 |
| Prothrombin | 1.187 | 1.041 | 1.353 | 1.75E-02 |
| Factor XI | 1.078 | 1.019 | 1.141 | 1.75E-02 |
| Factor XII | 1.047 | 1.009 | 1.087 | 1.82E-02 |
| Factor XIII | 1.033 | 1.002 | 1.065 | 3.80E-02 |
| **Healthy vs Moderate** |  |  |  |  |
| Antitrombin | 1.103 | 1.044 | 1.165 | 7.55E-04 |
| Prothrombin | 1.068 | 1.029 | 1.109 | 7.55E-04 |
| Factor XI | 1.056 | 1.024 | 1.088 | 7.55E-04 |
| Factor XII | 1.027 | 1.006 | 1.048 | 9.92E-03 |
| Factor XIII | 1.037 | 1.016 | 1.058 | 7.55E-04 |
| **Healthy vs Severe** |  |  |  |  |
| Antitrombin | 1.056 | 1.007 | 1.107 | 4.00E-02 |
| Prothrombin | 1.054 | 1.011 | 1.099 | 4.00E-02 |
| Factor XI | 1.028 | 1.005 | 1.052 | 4.00E-02 |
| Factor XII | 1.009 | 0.989 | 1.030 | 3.94E-01 |
| Factor XIII | 1.016 | 0.997 | 1.036 | 1.19E-01 |
| **AM vs Moderate** |  |  |  |  |
| Antitrombin | 1.003 | 0.971 | 1.036 | 8.54E-01 |
| Prothrombin | 0.991 | 0.972 | 1.010 | 5.60E-01 |
| Factor XI | 1.010 | 0.991 | 1.030 | 5.60E-01 |
| Factor XII | 0.996 | 0.984 | 1.007 | 5.60E-01 |
| Factor XIII | 1.014 | 0.997 | 1.031 | 5.60E-01 |
| **AM vs Severe** |  |  |  |  |
| Antitrombin | 0.991 | 0.957 | 1.025 | 8.47E-01 |
| Prothrombin | 0.998 | 0.989 | 1.007 | 8.47E-01 |
| Factor XI | 0.996 | 0.977 | 1.015 | 8.47E-01 |
| Factor XII | 0.973 | 0.953 | 0.994 | 6.15E-02 |
| Factor XIII | 0.999 | 0.981 | 1.017 | 8.87E-01 |
| **Moderate vs Severe** |  |  |  |  |
| Antitrombin | 0.980 | 0.960 | 1.000 | 6.16E-02 |
| Prothrombin | 1.001 | 0.995 | 1.008 | 7.24E-01 |
| Factor XI | 0.987 | 0.976 | 0.998 | 2.85E-02 |
| Factor XII | 0.987 | 0.976 | 0.997 | 2.85E-02 |
| Factor XIII | 0.982 | 0.972 | 0.993 | 5.01E-03 |

**Statistics:** Pairwise logistic regression models were fit for each protein and pair of COVID-19 severity classes. Age and sex were added in each model as covariables. Odds ratio, 95% confidence intervals (lower boundary: 2.5%, upper boundary: 97.5%), and the False Discovery Rate (FDR) q-values are shown for every model.

**Supplementary Table 4.** Survival analysis in COVID-19 patients according to coagulation proteins’
s levels.

| Cox<br>Proportional-Hazard | Cox Proportional-Hazard |  |  | Aalen's Additive<br>Regression |  |
| --- | --- | --- | --- | --- | --- |
|  | HR | p-value | C-index | Coefficient | p-value |
| **Antithrombin** |  |  | 0.865 ±0.035 |  |  |
| Antithrombin | 0.959 | 1.38E-03 |  | -1.84E-02 | 1.17E-03 |
| Age | 1.091 | 2.23E-05 |  | 6.12E-04 | 5.64E-05 |
| Sex | 2.282 | 1.43E-01 |  | 7.87E-03 | 9.12E-03 |
| **Prothrombin** |  |  | 0.819 ±0.056 |  |  |
| Prothrombin | 1.005 | 9.43E-02 |  | 5.62E-05 | 7.23E-01 |
| Age | 1.106 | 1.17E-06 |  | 5.85E-05 | 7.01E-05 |
| Sex | 3.273 | 3.06E-02 |  | 6.91E-03 | 1.05E-01 |
| **Factor_XI** |  |  | 0.832 ±0.045 |  |  |
| Factor_XI | 0.987 | 1.77E-01 |  | -1.25E-04 | 5.81E-03 |
| Age | 1.091 | 1.87E-05 |  | 6.24E-04 | 5.41E-04 |
| Sex | 2.692 | 7.41E-03 |  | 7.21E-03 | 7.74E-03 |
| **Factor_XII** |  |  | 0.851 ±0.043 |  |  |
| Factor_XII | 0.981 | 3.36E-03 |  | -8.92E-05 | 8.52E-03 |
| Age | 1.085 | 3.73E-05 |  | 5.65E-04 | 7.97E-04 |
| Sex | 2.312 | 1.31E-01 |  | 6.28E-03 | 1.48E-01 |
| **Factor_XIII** |  |  | 0.851 ±0.044 |  |  |
| Factor_XIII | 0.971 | 2.34E-03 |  | -1.28E-03 | 2.61E-04 |
| Age | 1.098 | 7.07E-06 |  | 6.34E-04 | 5.47E-04 |
| Sex | 2.141 | 1.89E-02 |  | 5.91E-03 | 1.56E-01 |

**Statistics:** Cox Proportional-Hazard and Aalen's Additive Regression. Hazard ratio, p-values and Harrell's concordance index (C-index) are shown for the Cox models. C-index is a goodness of fit measure for models that produces risk scores. Models with higher C-index indicate a shorter time-to-disease for those patients with higher risk score. A C-index's value of 0.5 entails that the risk score predictions are no better than chance. Values near 1 indicates perfect separation of patients with different outcomes. Regression coefficients and their associated p-values are also shown for the Aalen's models. The Aalen model allows for time-varying covariate effects. Regarding the covariables it is possible to notice that age and sex have a positive and significant effect over mortality. Abbreviations: HR, hazard ratio.

**Supplementary Figure 1.**
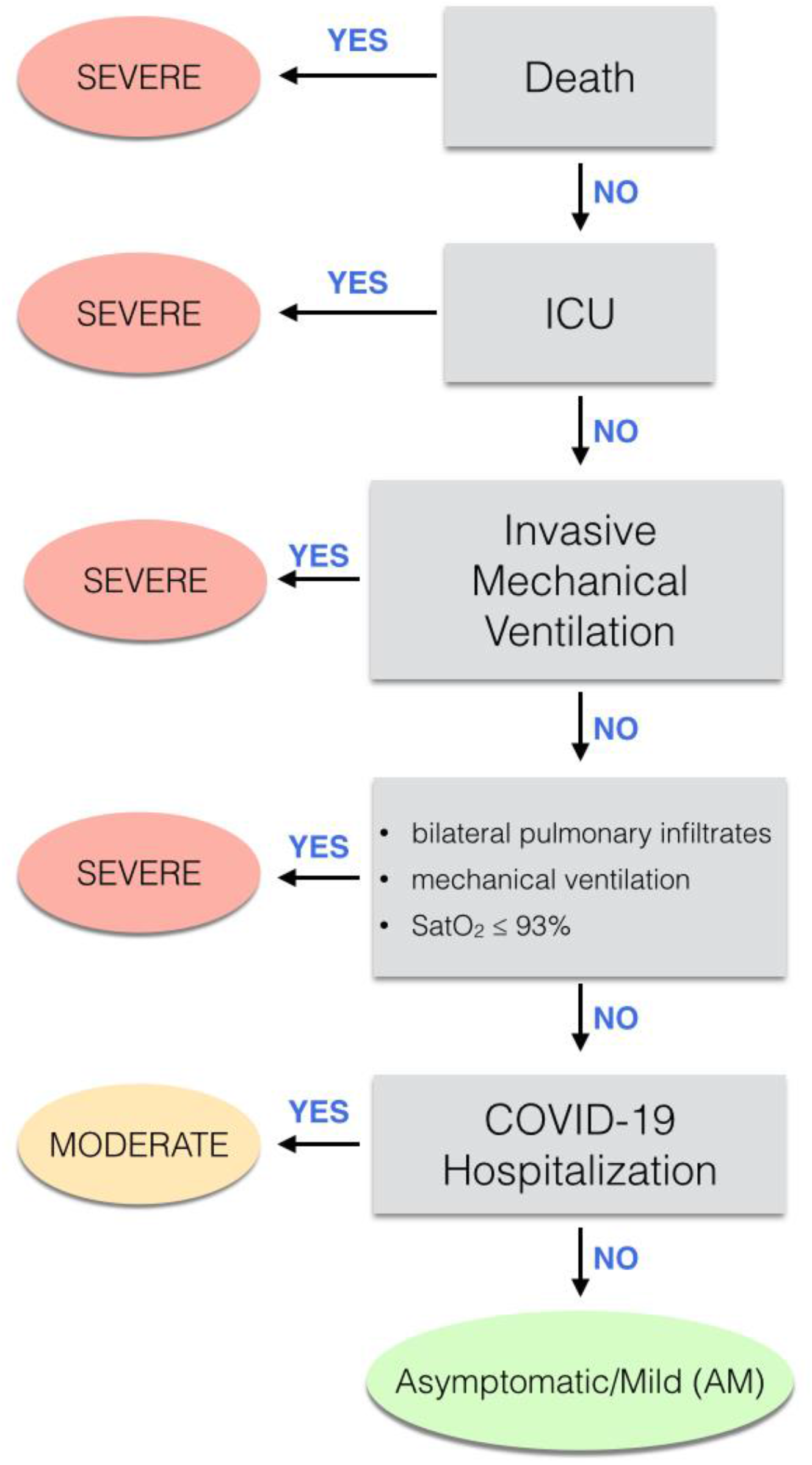
Flowchart describing the classification of patients with respect to COVID-19 severity. **Abbreviations:** ICU. Intensive care unit; ARDS, acute respiratory distress syndrome; AM: asymptomatic/mild patients.

**Supplementary Figure 2.**
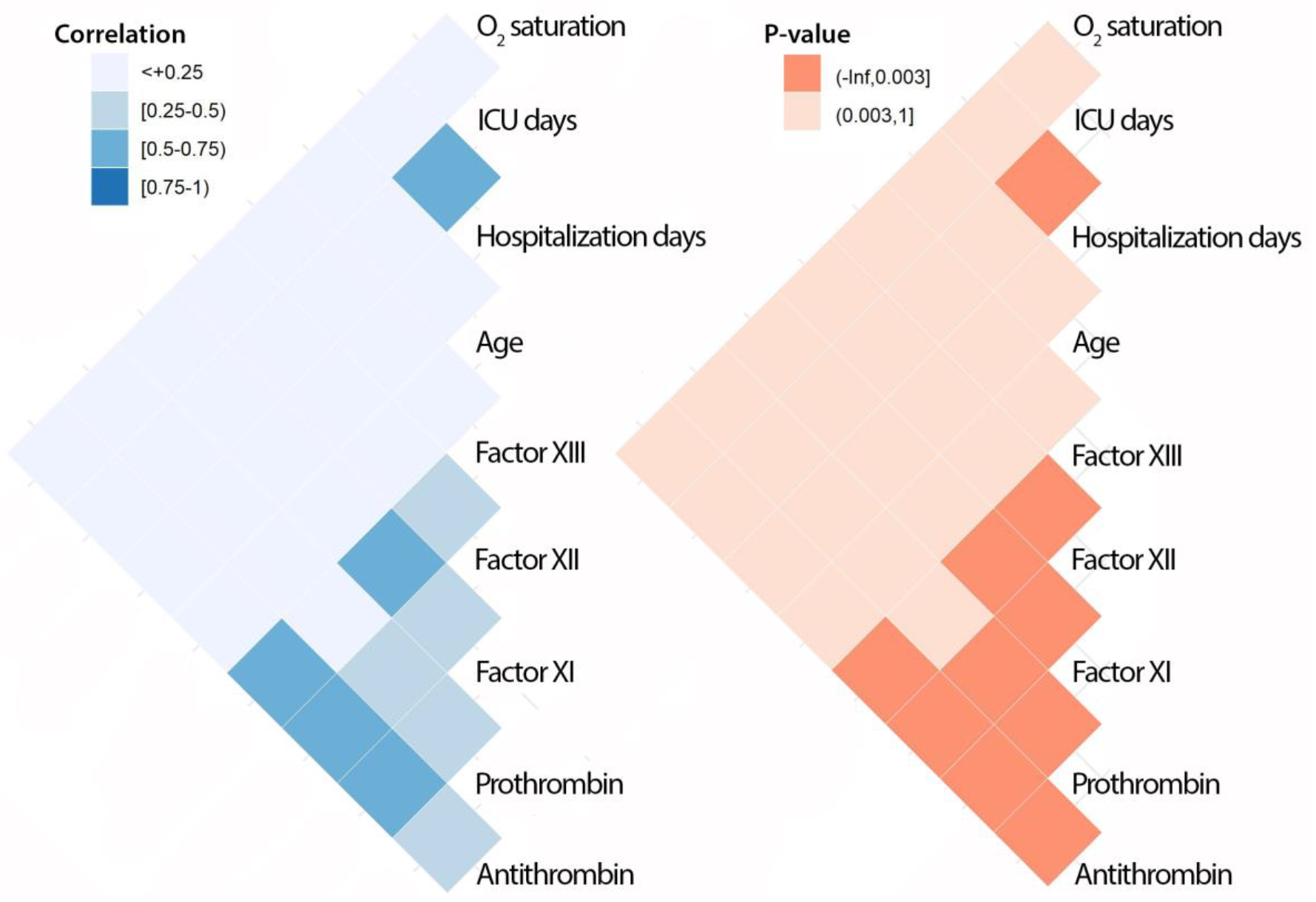
Correlation between coagulation proteins and continuous covariables related to severity. **Statistics:** Pearson correlation. Heatmaps are provided for the correlation coefficients and its associated p-values. A regular Bonferroni correction for multiple testing was applied, being significance set at 3.0e^-03^. **Abbreviations**: O_2_, oxygen; ICU, intensive care unit.

**Supplementary Figure 3.**
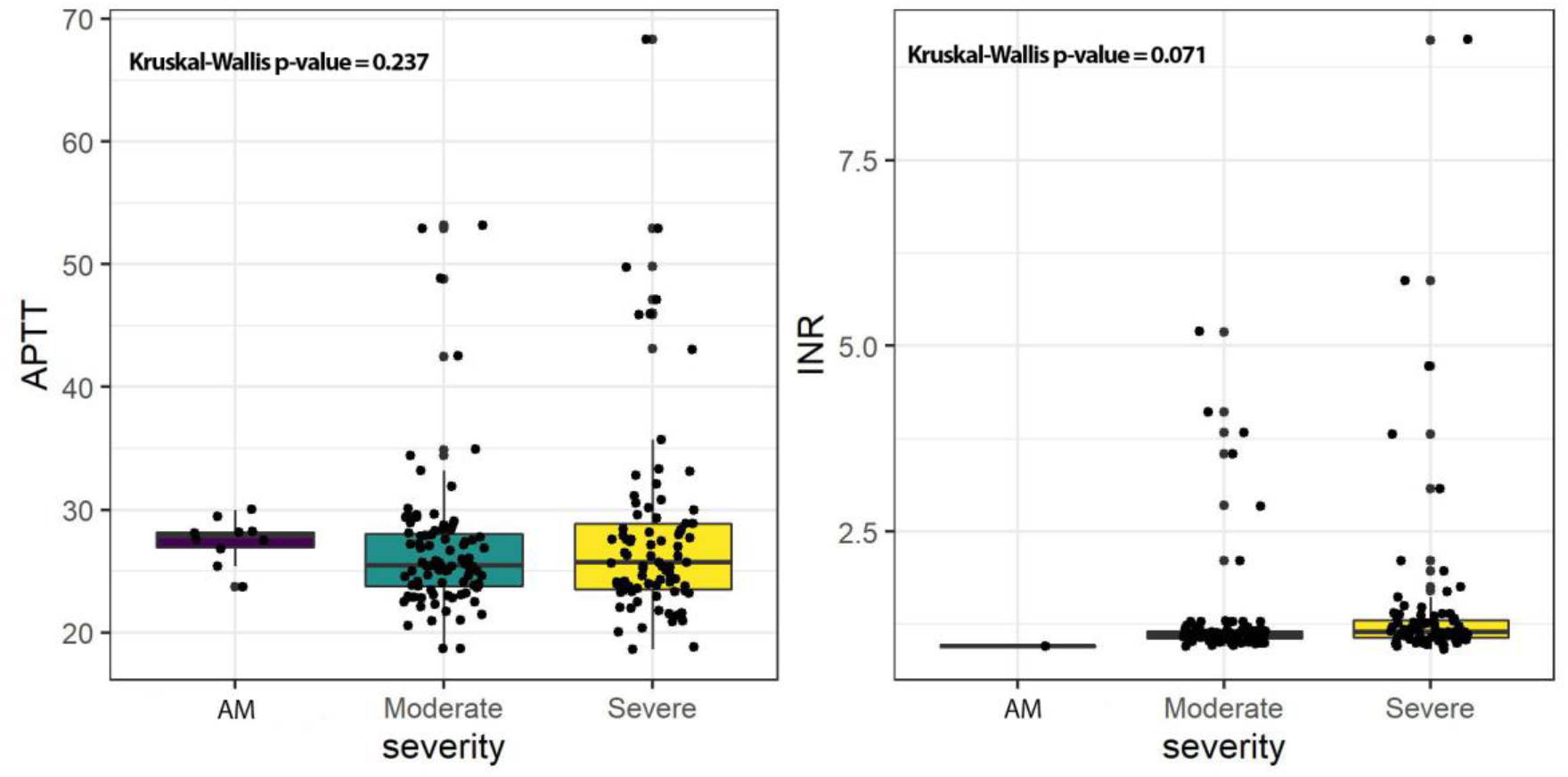
Differences in activated partial thromboplastin time (APTT) and international normalized ratio (INR) between the severity groups. **Statistics:** Distributions were presented in box plots and significance was calculated by Kruskal-Wallis test. APTT and INR levels did not show differences among the three groups of COVID-19 (p-value = 0.237 and p-value = 0.071, respectively). **Abbreviations**: AM, asymptomatic/mild patients; APTT, activated partial thromboplastin time; INR, international normalized ratio.

**Supplementary Figure 4.**
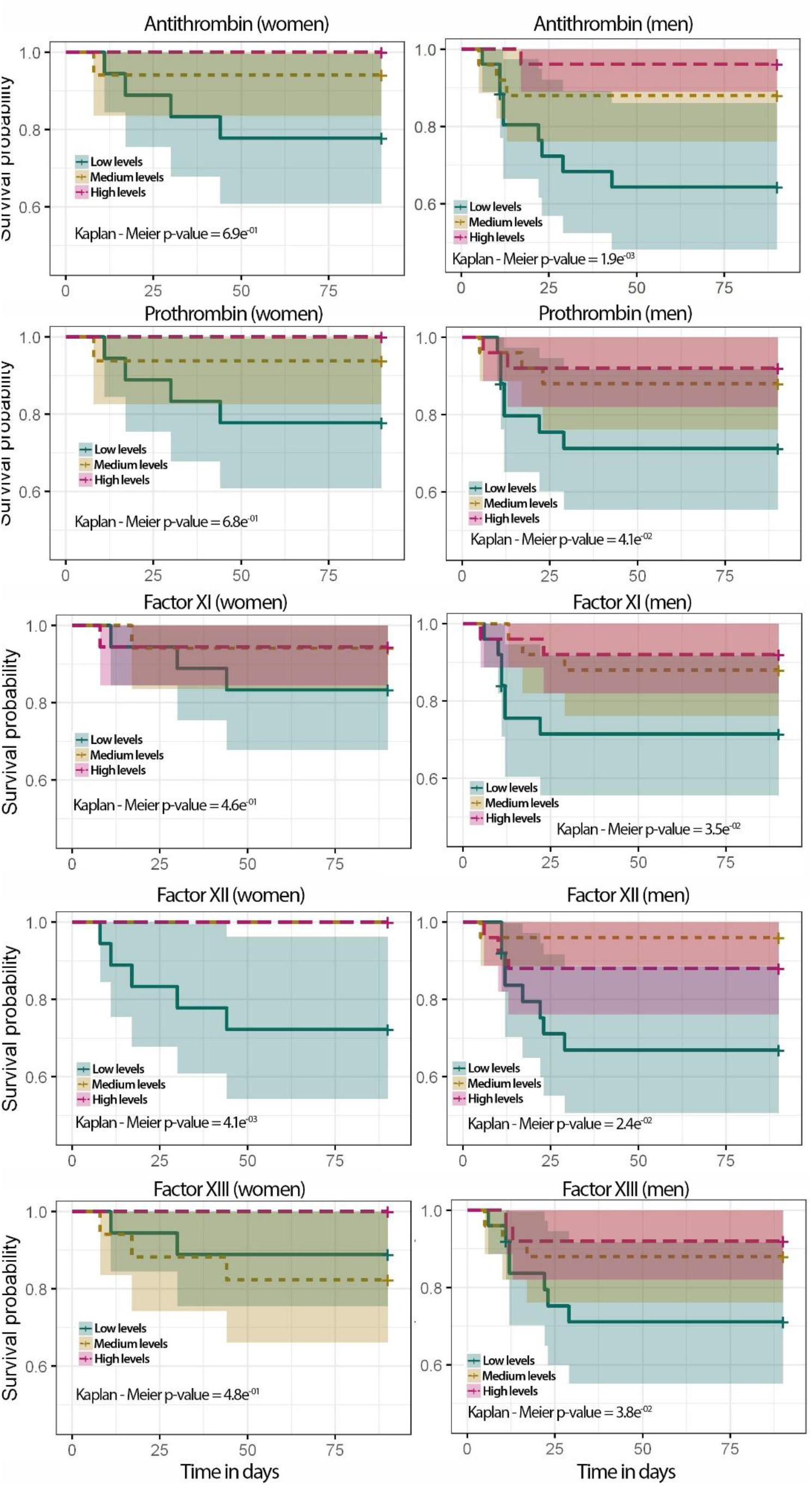
Kaplan-Meier plots regarding coagulation protein levels and grouped by sex. **Statistics**: Coagulation proteins’
s classes were obtained using 3 quantiles to get low (blue), medium (yellow) and high (pink) factor levels. P-values of the Kaplan-Meier analysis are shown in each plot. Men = 75, Women = 53.

**Supplementary Figure 5.**
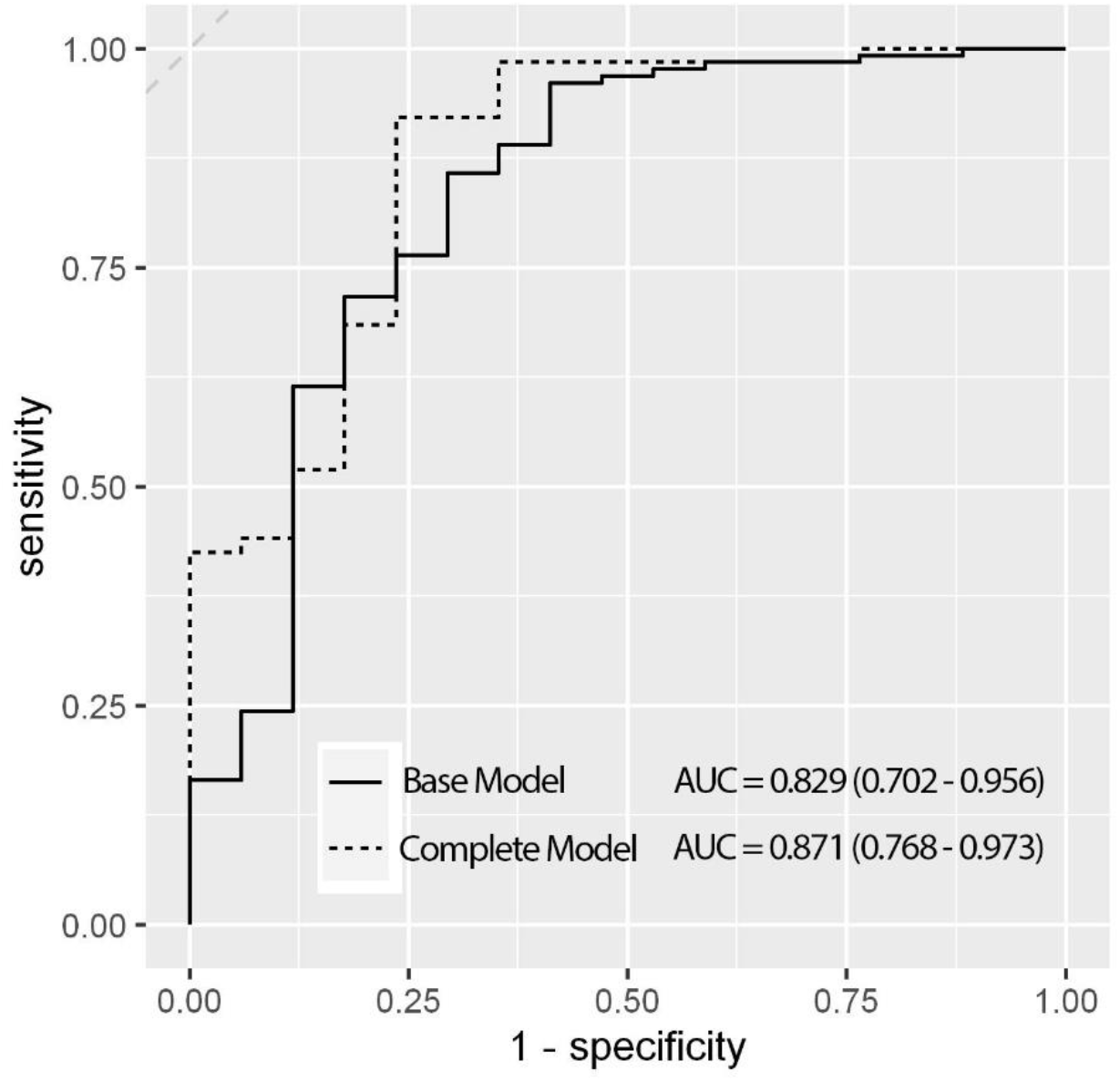
Predictive accuracy of the model with coagulation proteins in combination with epidemiological variables. **Statistics:** ROC curves and the Area Under the Curve (AUC) with its 95% CI in brackets. Base model: mortality is modelled according to age and sex covariables. Complete Model: all five coagulation protein levels were included (antithrombin, prothrombin, Factor XI, Factor XII, Factor XIII) to the base model. No significant differences were found between models.

